# Unwelcome memento mori or best clinical practice? Community end-of-life anticipatory medication prescribing practice: a mixed methods observational study

**DOI:** 10.1101/2021.02.25.21252474

**Authors:** Ben Bowers, Kristian Pollock, Stephen Barclay

## Abstract

**Background:** Anticipatory medications are injectable drugs prescribed ahead of possible need for administration if distressing symptoms arise in the final days of life. Little is known about how they are prescribed in primary care.

**Aim:** To investigate the frequency, timing and recorded circumstances of anticipatory medications prescribing for patients living at home and in residential care.

**Design:** Retrospective mixed methods observational study using General Practitioner and community nursing clinical records.

**Setting/participants:** 329 deceased adult patients registered with Eleven General Practitioner practices and two associated community nursing services in Hertfordshire and Cambridgeshire, England (30 most recent deaths per practice). Patients died from any cause except trauma, sudden death or suicide, between 4 March 2017 and 25 September 2019.

**Results:** Anticipatory medications were prescribed for 167/329 (50.8%) of the deceased patients, between 0 and 1212 days before death (median 17 days). The likelihood of prescribing was significantly higher for patients with a recorded preferred place of death (odds ratio [OR] 34; 95% CI 15-77; *p* < 0.001) and specialist palliative care involvement (OR 7; 95% CI 3-19; *p* < 0.001). For 66.5% of patients (111/167) anticipatory medications were recorded as being prescribed as part of a single end of-life planning intervention.

**Conclusion:** The variability in the timing of prescriptions highlights the challenges in diagnosing the end-of-life phase and the potential risks of prescribing far in advance of possible need. Patient and family preferences for involvement in anticipatory medications prescribing decision-making and their experiences of care warrant urgent investigation.

**Summary Box:** What is already known on this topic

1. The prescribing of injectable anticipatory medications to provide symptom relief in the last days of life care is recommended and widespread practice in a number of counties.
2. There is limited research concerning the frequency, timing and context of prescriptions.

What this paper adds

1. Half (50.8%) of 319 patients whose deaths were potentially predictable deaths were prescribed anticipatory medications, the timing of prescriptions ranging from 0 to 1212 days before death (median 17 days).
2. Anticipatory medications were frequently prescribed as standardised drugs and doses, and often as part of a single end-of-life care planning intervention.
3. The extent to which patients and family carers were involved in prescribing decisions was unclear.

Implications for practice, theory or policy

1. Patient and family preferences for involvement in anticipatory medications prescribing decision-making and their experiences of care warrant urgent investigation.
2. The presence of anticipatory medications for long periods of time may compromise patient safety unless robust systems are in place to review their continued appropriateness and safe use.

## Introduction

Timely and effective symptom control in the last days of life is a key component in ensuring a comfortable death. (1-6) In the UK, Australia, Canada, Norway and New Zealand, the individualised prescribing of injectable anticipatory medications (AMs) for people approaching the end of life in the community ahead of potential need is widely promoted to optimise symptom control in the last days of life at home and prevent unwanted hospital admissions. (7-13) AMs are kept in the home, where they are available to be administered by visiting nurses or doctors if the patient is unable to take oral medications and develops symptoms of pain, breathlessness, nausea and vomiting, agitation or respiratory secretions at the end of life. (7,14,15)

Although anticipatory prescribing is recommended practice in several countries (8-9,12-13) there is inadequate evidence of its clinical effectiveness and limited research into the incidence and timing of prescriptions. (7,8) Reported prescribing rates vary from 16% of predictable death in a primary care population (16) to 63% of patients receiving specialist palliative care input. (17) Patients with advanced cancer appear more likely to receive prescriptions than those with non-cancer terminal conditions. (11,18-20) The timing of prescribing to death is reported as ranging from a few days, (11,19,20) several weeks (15,17,19-21) to several months before death. (22)

The decision to prescribe AMs is multifaceted and little studied to date. Community nurses report they initiate anticipatory prescribing conversations with patients and families when they perceive that death is imminent, following which they prompt General Practitioners (GPs) to prescribe AMs. (15,23,24) Nurses find some GPs to be resistant to prescribing AMs (18,23,25) while other GPs act on their requests to prescribe. (23,24) Some GPs prefer to independently judge when to prescribe AMs, (20,22) or to discuss intended care with patients themselves and prescribe AMs whilst their condition is stable. (22) Prescribing decisions involve assessing patient and family willingness to have end-of-life care discussions, safety risks associated with prescribing strong opioids and how soon medications may be needed. (15,20,22)

GP and community nurse records provide useful observational data for understanding practice. Retrospective examination of routinely collected clinical data enables investigation of recorded activities and interactions such as prescribing while avoiding selection and recruitment biases that are a major difficulty in prospective studies of terminally ill populations. (26,27) Our study aims were to investigate the frequency, timing and recorded circumstances of injectable end-of-life anticipatory medications (AMs) prescribing for patients living at home and in residential care.

## Methods

### Study design

We carried out a retrospective mixed methods observational study of deceased patients GP and community nursing held clinical records. (28,29) Reflecting the social constructionist paradigm, (30) clinical records are selective and stylised clinician accounts rather than presenting only objective facts. (31,32)

The Cambridge Positive Ageing and Cambridge Palliative and End of Life Care Patient and Public Involvement (PPI) Groups supported the study throughout, advising on research priorities, the acceptability of accessing deceased patient records without consent and interpretation of key findings. The South Cambridgeshire Research Ethics Committee granted ethics approval [Reference: 19/EE/0012]. The Health Research Authority’s Confidentiality Advisory Group [19/CAG/0014] approved the processing of confidential patient information without consent in the public interest: data were anonymised at the earliest opportunity.

### Study population and setting

Participants were registered with eleven GP practices and two associated National Health Service Community Trusts providing community nursing services in Hertfordshire and Cambridgeshire. GP practices were purposively sampled from 21 practices expressing interest in participation, to obtain a maximum diversity sample in terms of patient list sizes (range 5,500 to 43,000), geographical setting (two outer London practices, three urban, six rural town / villages) and socioeconomic status (range third most deprived decile to the least deprived decile).

Each practice identified the 30 most recent expected deaths of patients aged >17 years, who had been living at home or in residential care for at least one day in the last month of life and had died from any cause except trauma, sudden death or suicide. Patients who had previously indicated a wish not to be involved in research were excluded. Patients died between 4 March 2017 and 25 September 2019: one was excluded upon confirming their cause of death after data extraction, leaving a study population of 329 patients.

### Data sources and definitions

The electronic GP records and electronic and paper community nursing records of the deceased patients were retrieved and examined between May 2019 and March 2020 by BB, an experienced community and palliative care nurse. Patient demographic and clinical characteristics, documented end-of-life planning discussions and decisions, summary of events in the seven days preceding AM prescribing, recorded prescribing contexts and decision-making, and medication details were entered into a custom-built secure database (Supplemental Document 1). Relevant free text record entries were summarised. Cause and date of death were confirmed from GP practice held death certificate books or England’s General Register Office.

AMs were defined as one or more injectable medications prescribed ahead of need to be administered for symptom control in the last days of life. (7,14)

### Data analysis

Data analysis combined quantitative and qualitative analyses in a mixed methods approach. (28,29) Categorical data are reported as frequencies and percentages and continuous data as median (interquartile range: IQR). The sample size of 330 patients was calculated a priori with a statistician to enable statistical analysis including Chi-square and Fisher’s exact test and multivariable logistic regression models. (29). Data analysis was performed using SPSS version 26: P<0.05 is considered statistically significant.

BB undertook qualitative analysis using inductive constant comparison incident-to-incident coding (28) of extracted data from the clinical records of patients prescribed AMs, focusing on end-of-life discussions, AM prescribing contexts and associated patient and family interactions, using NVivo version 12. Patterns and variances in records, typologies of care, and decisions in attributing significance to findings were discussed and refined with KP, SB and both PPI groups. These iterative steps informed the interpretative analysis. (28,33)

## Results

Most deceased patients were either aged between 75 and 84 years (92/329, 28%) or 85 years and older (124/329, 37.7%). The majority of deaths were from non-cancer conditions (193/329, 58.7%). See **Table 1**.

**Table 1.** Demographics and clinical characteristics of deceased patients.

| **Age range** | **n** | **(%)** |
| --- | --- | --- |
| 18-64 | 50 | (15.2%) |
| 65-74 | 63 | (19.1%) |
| 75-84 | 92 | (28.0%) |
| 85+ | 124 | (37.7%) |
| **Gender male** | 169 | (51.4%) |
| **Usual place of care** |  |  |
| Home | 299 | (90.9%) |
| Care Home | 30 | (9.1%) |
| **Ethnicity** |  |  |
| White | 294 | (89.4%) |
| Other | 11 | (3.3%) |
| Not recorded | 24 | (7.3%) |
| **Cause of death** |  |  |
| Cancer: solid tumour | 130 | (39.5%) |
| Cancer: haematological malignancy | 6 | (1.8%) |
| Chronic heart disease | 41 | (12.5%) |
| Dementia | 15 | (4.6%) |
| Pneumonia | 48 | (14.6%) |
| Chronic obstructive pulmonary disease | 26 | (7.9%) |
| Stroke | 13 | (4%) |
| Liver disease | 2 | (0.6%) |
| Acute heart disease | 3 | (0.9%) |
| Frailty of old age | 22 | (6.7%) |
| Other | 23 | (7%) |

In total, 167/329 (50.8%) patients were prescribed AMs. There was a wide range of prescribing rates across the eleven GP practices, with a median of 14 / 30 patients (46.7%) (IQR 11 – 17 / 30 patients) and range 7 / 30 (23.3%) to 28/30 (93.3%). There was a highly statistically significant association between the GP practice patients were registered with and whether they were prescribed AMs (p<0,001). Patients who died from cancer were more likely to be prescribed AMs (67.6%) than those who died from non-cancer conditions (38.9%) (p < 0.001). See **Table 2**.

**Table 2.** Univariate analysis of relationships of patient characteristics and anticipatory medication prescribing.

| Patient characteristics | Prescribed anticipatory medications (167) / Total (329) |  |  |
| --- | --- | --- | --- |
| **Gender** | | $X^2$ (DF = 1) = 0.503 | $p = 0.478$ |
| Male | 89/169 (52.7%) |  |  |
| Female | 78/160 (48.8%) |  |  |
| **Age Range** | | $X^2$ (DF = 3) = 1.345 | $p = 0.718$ |
| 18-64 | 24/50 (48%) |  |  |
| 65-74 | 33/63 (52.4%) |  |  |
| 75-84 | 43/92 (46.7%) |  |  |
| 85+ | 67/124 (54%) |  |  |
| **Ethnicity** | | $X^2$ (DF = 2) = 0.304 | $p = 0.859$ |
| White | 150/294 (51%) |  |  |
| Other | 6/11 (54.5%) |  |  |
| Not Recorded | 11/24 (45.8%) |  |  |
| **GP Practice ID No.** | | $X^2$ (DF = 10) = 36.059 | $p < 0.001$ |
| One | 13/30 (43.3%) |  |  |
| Two | 14/30 (46.7%) |  |  |
| Three | 14/30 (46.7%) |  |  |
| Four | 28/30 (93.3%) |  |  |
| Five | 19/29 (65.5%) |  |  |
| Six | 16/30 (53.3%) |  |  |
| Seven | 16/30 (53.3%) |  |  |
| Eight | 14/30 (46.7%) |  |  |
| Nine | 13/30 (43.3%) |  |  |
| Ten | 7/30 (23.3%) |  |  |
| Eleven | 13/30 (43.3%) |  |  |
| **Number of practice chronic disease registers patient was on** | | $\chi^2$ (DF = 4) = 12.789 | $p = 0.012$ |
| 0-1 | 12/43 (27.9%) |  |  |
| 2-3 | 64/128 (50%) |  |  |
| 4-5 | 48/88 (54.5%) |  |  |
| 6-7 | 27/43 (62.8%) |  |  |
| 8-13 | 16/27 (59.3%) |  |  |
| **Usual Place of Residence** | | $\chi^2$ (DF = 1) = 0.728 | $p = 0.393$ |
| Care Home | 13/30 (43.3%) |  |  |
| Home | 154/299 (51.5%) |  |  |
| **Cause of Death** | | $\chi^2$ (DF = 1) = 26.452 | $p < 0.001$ |
| Cancer | 92/136 (67.6%) |  |  |
| Non-Cancer | 75/193 (38.9%) |  |  |
| **DNACPR form completed** | | Fisher's Exact Test | $p < 0.001$ |
| Yes | 163/227 (71.8%) |  |  |
| No | 4/102 (3.9%) |  |  |
| **On GP Practice Palliative Care Register** | | $\chi^2$ (DF = 1) = 120.321 | $p < 0.001$ |
| Yes | 135/168 (80.4%) |  |  |
| No | 32/161 (19.9%) |  |  |
| **Received Specialist Palliative Care** | | $\chi^2$ (DF = 1) = 86.166 | $p < 0.001$ |
| Yes | 117/148 (79.1%) |  |  |
| No | 50/181 (27.6%) |  |  |
| **Preferred Place of Death** | | $\chi^2$ (DF = 1) = 186.118 | $p < 0.001$ |
| Recorded | 152/178 (85.4%) |  |  |
| Not Recorded | 15/151 (9.9%) |  |  |

All statistically significant variables in the univariate analysis in Table 2 were entered into a multivariate regression analysis, which revealed that after adjustment for gender, age range, GP practice, number of chronic disease registers on, usual residence and cause of death, the likelihood of being prescribed AMs was significantly higher for patients with a recorded preferred place of death (OR 34; 95% CI 15-77; *p* < 0.001) and for patients who had received specialist palliative care (OR 7; 95% CI 3-19; *p* < 0.001) (Supplementary Document 2). Preferred place of death was included in the logistic regression model as the most theoretically sound marker of end-of-life planning. A completed Do Not attempt Cardiopulmonary Resuscitation (DNACPR) form and / or inclusion on the GP practice palliative care register was not always associated with a record of explicit end-of-life care planning.

### Recorded prescribing decision-making

AMs were frequently recorded as being be prescribed as part of a single end of-life planning consultation: for 111/167 (66.5%) of patients prescribed AMs, the prescription (or discussion concerning prescription) occurred during the same consultation when preferred place of death and / or cardiopulmonary resuscitation discussions were first recorded. For 78/167 (46.7%) of patients prescribed AMs, this was when the end of life appeared imminent, with rapid deterioration of physical strength over a few days, escalating symptoms, and reduced ability to eat or drink. In contrast, for 72/167 patients (43.1%), AMs were prescribed as part of longer-term forward planning, when patients had relatively stable physical function, but the focus of care had shifted to end-of-life support. For a further 17/167 (10.2%), AMs were prescribed in the context of clinical uncertainty, when oral antibiotics were prescribed for potentially reversible infections alongside AMs if their condition did not improve:

> *6 days before death, GP visits the patient and records: “Deterioration, taken to bed, refusing drinks for the last 1-2 days. Comfortable, responding to carers*… *Unclear if has a urine tract infection (UTI) or this is a pre-terminal event… Tried phoning family but no answer. Plan: Completed DNACPR form with agreement of carers. Issued anticipatory medications and oral antibiotics. Treat for UTI and encourage fluids*.*” Computer codes: preferred place of care and death is home, patient is ‘aware of prognosis’. [Patient 04014C]*

Recorded patient and family involvement in decisions to prescribe AMs were variable: no prescribing conversations were recorded for 69/167 of patients (41.3%). AMs were typically prescribed following consultations involving conversations with patients about their preferences for having end-of-life care at home or their agreement with DNACPR decisions. For a few patients (6/167, 3.6%), it was recorded that they did not want to discuss their prognosis or consider that they were dying at the time of prescribing AMs. These patients lived alone, and prescribing decisions were framed as being in their best interests:

> *27 days before death, GP visits patient and records: Three recent hospital admissions in the last month for congestive cardiac failure symptom management. “Can barely get out of bed and needing carer visits four times a day… Does not want to discuss their prognosis… States wants active treatment and not wanting to engage in end-of-life care planning… Lives alone … May need to go into palliative care mode swiftly. Plan: best set [prescribe] anticipatory medications now*.*” Computer codes: preferred place of care and death is home. [Patient 01024D]*

Patient and family involvement in decision-making was recorded for 71/167 (42.5%) of patients, the records focussing on whether they agreed with clinician decisions to prescribe AMs: 10/71 patients (6%) were prescribed AMs prior to a visit or phone call to discuss prescribing. Most records concerning AM prescribing conversations were very brief, largely limited to reporting that families had been asked to collect the medications or patient / family agreement with prescribing decisions.

More detailed AM decision-making conversations were recorded when patients or families were concerned about possible symptoms (29/167, 17.4%). In a few cases (5/167, 3%), patient or families were recorded as not agreeing with a decision to prescribe AMs: three patients and one family were resistant to the idea of prescribing AMs, and one patient was “aggrieved” on discovering they had been prescribed AMs without being asked. In these cases, clinicians recorded that it was in the patient’s best interests to have medications available and documented persuading them to accept prescriptions:

> *3 days before death, GP registrar visits patient at home: “Discussed DNACPR and patient does not want resuscitation. States would like to pass away peacefully… Discussed anticipatory medications. [Patient states] does not want or need any medications. Explained these medications were only for if in distress… [Patient] remained adamant that does not need them… Discussed each group of medication and intended benefit in detail… I advised that they might not need them, and we will of course adhere to their wishes, but we do not want them to suffer… Agreed to [having] them*.*” [Patient 04025K]*

### Timing of prescribing

AMs were prescribed between 0 and 1212 days before death. Patients who died from cancer were prescribed AMs a median of 21.5 days before death (IQR 7 to 42 days, range 0 to 375 days); for those who died from non-cancer illnesses, AMs were prescribed a median of 12 days before death (IQR 4 to 47 days, range 1 to 1212 days). Seven patients were prescribed AMs a year or more before death, of which six had a non-cancer diagnosis. The median prescribing timing was 17 days before death across the eleven GP practices, with range of median of 6 to 33 days between the practices. See **Table 3**.

**Table 3.**
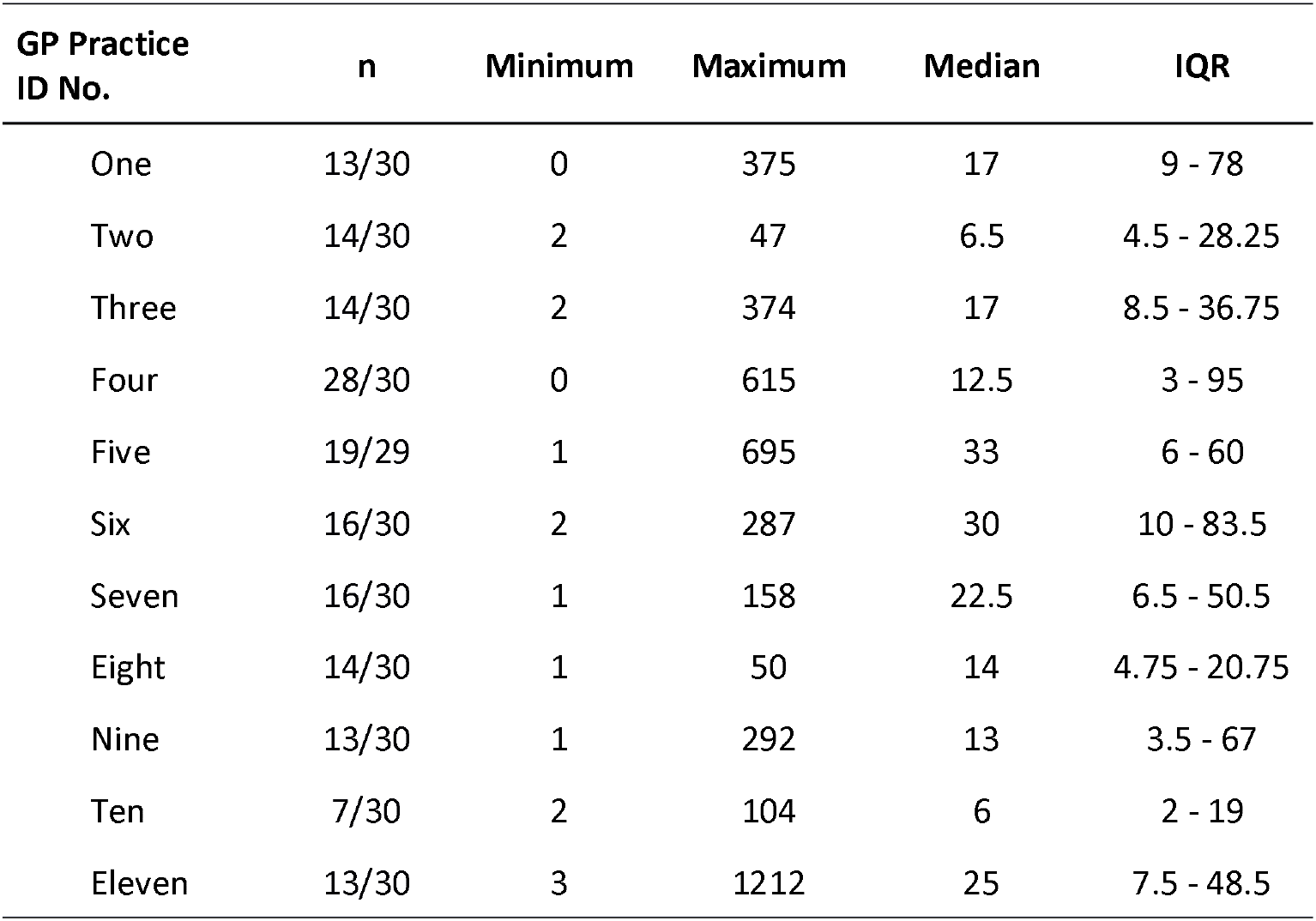
Timing of anticipatory medications prescribing in days before death.

| GP Practice ID No. | n | Minimum | Maximum | Median | IQR |
| --- | --- | --- | --- | --- | --- |
| One | 13/30 | 0 | 375 | 17 | 9 - 78 |
| Two | 14/30 | 2 | 47 | 6.5 | 4.5 - 28.25 |
| Three | 14/30 | 2 | 374 | 17 | 8.5 - 36.75 |
| Four | 28/30 | 0 | 615 | 12.5 | 3 - 95 |
| Five | 19/29 | 1 | 695 | 33 | 6 - 60 |
| Six | 16/30 | 2 | 287 | 30 | 10 - 83.5 |
| Seven | 16/30 | 1 | 158 | 22.5 | 6.5 - 50.5 |
| Eight | 14/30 | 1 | 50 | 14 | 4.75 - 20.75 |
| Nine | 13/30 | 1 | 292 | 13 | 3.5 - 67 |
| Ten | 7/30 | 2 | 104 | 6 | 2 - 19 |
| Eleven | 13/30 | 3 | 1212 | 25 | 7.5 - 48.5 |

### Clinicians prescribing medications

For 71/167 (42.5%) of patients issued AMs, requests for prescriptions came from clinicians different to the prescriber: specialist palliative care team members (42/167, 25.1%), community nurses (20/167, 12%); GP practice-based paramedics (3/167, 1.8%); care home staff (2/167, 1.2%). GPs in all eleven GP practices (37/167, 22.2%) prescribed AMs following requests from palliative care specialist or community nurse colleagues without recorded contact with the patient or family.

The majority of AMs (127/167, 76%) were prescribed by GPs: other prescribers included hospital doctors (25/167, 15%), nurse prescribers (7/167, 4.2%), out of hours doctors (6/167, 3.6%) and specialist palliative care doctors (2/167, 1.2%).

### Symptom control prescribing

The majority of patients (154/167, 92.2%) were prescribed AMs for all five common end-of-life symptoms: pain, breathlessness, nausea and vomiting, agitation and respiratory tract secretions. Similar drugs and dose ranges were prescribed for all five symptoms following end-of-life electronic record template recommendations for 105/167 (62.9%) of patients. See **Table 4**.

**Table 4.** Anticipatory medications prescribed.

| Drug name | n | Percentage |
| --- | --- | --- |
| **Opioid** | **165/167** | **98.8%** |
| Morphine Sulfate <sup>2</sup> | 114/167 | 68.3% |
| Oxycodone <sup>2</sup> | 26/167 | 15.6% |
| Diamorphine <sup>2</sup> | 25/167 | 15.0% |
| **Anxiolytic** | **166/167** | **99.4%** |
| Midazolam <sup>1</sup> | 166/167 | 99.4% |
| **Anti-emetic** | **159/167** | **95.2%** |
| Haloperidol <sup>3</sup> | 97/167 | 58.0% |
| Cyclizine <sup>3</sup> | 54/167 | 32.3% |
| Levomepromazine <sup>3</sup> | 5/167 | 3.0% |
| Metoclopramide <sup>3</sup> | 2/167 | 1.2% |
| Ondansetron <sup>3</sup> | 1/167 | 0.6% |
| **Antisecretory** | **163/167** | **97.6%** |
| Glycopyrronium <sup>4</sup> | 152/167 | 91.0% |
| Hyoscine Butylbromide <sup>4</sup> | 9/167 | 5.4% |
| Hyoscine hydrobromide <sup>4</sup> | 2/167 | 1.2% |

Recorded reason for prescription:
<sup>1</sup> restlessness or agitation
<sup>2</sup> pain relief and shortness of breath
<sup>3</sup> nausea and vomiting
<sup>4</sup> respiratory tract secretions.

### Anticipatory syringe drivers

For 49/167 (29.3%) of patients, a prescription for a continuous subcutaneous infusion of end-of-life care drugs was also issued ahead of need. These “anticipatory syringe drivers” were usually for the same medications as AM injections, often with larger dose ranges. The frequency and timing of anticipatory syringe driver prescriptions varied widely between GP practices, ranging from 1/16 patients (6.3%) to 10/14 patients (71.4%), with prescribing timing a median of 5.5 days before death across the eleven GP practices (range 2 to 27 days). See **Table 5**.

**Table 5.** Timing of anticipatory syringe driver prescribing in days before death.

| GP Practice ID No. | n | Minimum | Maximum | Median | IQR |
| --- | --- | --- | --- | --- | --- |
| One | 3/13 | 6 | 94 | 27 | - |
| Two | 10/14 | 2 | 46 | 5.5 | 2.75 - 24.5 |
| Three | 7/14 | 1 | 18 | 2 | 2 - 9 |
| Four | 4/28 | 1 | 164 | 19.5 | 1.5 - 132 |
| Five | 4/19 | 1 | 36 | 2.5 | 1.25 - 27.75 |
| Six | 3/16 | 0 | 17 | 4 | - |
| Seven | 1/16 | 2 | 2 | 2 | - |
| Eight | 4/14 | 1 | 18 | 10.5 | 2 - 17.5 |
| Nine | 3/13 | 1 | 49 | 16 | - |
| Ten | 2/7 | 4 | 5 | 4.5 | - |
| Eleven | 8/13 | 2 | 536 | 15 | 3.5 - 102 |

## Discussion

Our study is the first to highlight the high frequency and standardised prescribing of AMs prescriptions for terminally ill patients in a primary care population. The range of timing of prescribing identified contrasts with the published evidence reporting that prescribing is limited to a few days to several weeks before death. (19,20) Our findings correspond with GPs’ and nurses’ accounts of preferring to put AMs in place as early as is feasible. (15,21,22)

Prognostication is a very inexact science. It is difficult to predict the timing of death (34-37) particularly for those with a highly unpredictable chronic frailty dying trajectory. (20,38,39) Although AMs are typically prescribed closer to death for patients with non-cancer conditions, (19) six of the seven patients in our study prescribed AMs a year or more before death had non-cancer diagnoses. Some patients were prescribed AMs alongside antibiotics when there was clinical uncertainty about whether they were dying or had reversible infections. The prescribing of AMs is a highly symbolic intervention and can be perceived by visiting clinicians who are unfamiliar with the patient as a signal of poor prognosis and imminence of dying, (22) even when this may not yet be the case.

Anticipatory syringe driver prescribing was common practice in several of the study GP practices. The recent Gosport War Memorial Hospital inquiry in the UK has highlighted the particular dangers for patient safety when prescribing anticipatory syringe drivers with large dose ranges to be started at the discretion of third parties whose clinical assessment skills are unknown to the prescriber. (7,40) There is no previously published research on the practice of prescribing anticipatory syringe drivers: research is urgently needed to investigate the clinical appropriateness and safety of anticipatory syringe driver prescribing. (41)

Palliative care teams often initiate end-of-life care planning interventions including AMs prescription requests. (15,17,42) Patients who had seen a specialist palliative care team were seven times more likely to be prescribed AMs. A referral to specialist palliative care, or the involvement of such a team, may again be perceived to be a signal to everyone involved that the patient is approaching end-of-life, which at times may not be the case.

End-of-life care planning is presented in current policy and clinical discourse as an evolving and individualised process that is started with patients whilst their condition is stable, with regular reviews as their situation and preferences change. (8,43-46) In keeping with previous research, we found advance care planning decisions were frequently recorded as part of one main end-of-life care consultation or crisis intervention that comprises identifying preferred place of death, putting in place AMs and completing a DNACPR form. (22,47,48) Primary care electronic end-of-life record templates, increasingly used in the UK, aid communication and continuity of care. (26,37,49) This technology also shapes practice and may inadvertently encourage the bureaucratization of end-of-life care planning interventions by promoting a ‘one size fits all’ process. (22,32,50,51) There is a tension between using end-of-life templates to provide standard guidance whilst promoting personalised care.

The preferences of clinicians and expectations of policymakers for ensuring that end-of-life advance care plans, including AMs, are in place, need to be balanced with patient and family readiness to have sensitive discussions and to make plans for future care. (8,44,47) Our analysis found clinical records were largely silent about conversations with patients and family members concerning the implications and emotional impacts of AM prescribing. Corresponding with previous research, there were occasions where professional led end-of-life planning, including the prescribing of AMs, took place without consultation with patients unwilling or unable to consider future care. (22,47,48) Patient and family preferences for involvement in AM prescribing decision-making and their experiences of care warrant urgent investigation. (7,22,52)

### Strengths and limitations

Caution is needed in interpreting what records can tell us about patient and family participation in prescribing decisions. Records only contain a small part of any clinical encounter, the emphasis frequently being on clinical decisions and prescribing matters. (32) The lack of recorded information on patient understanding of AMs and their preferences is problematic as records are considered authoritative and influence subsequent care decisions. (31,32)

The generalisability of the results is enhanced by the identification of sequential deaths and purposive sampling of GP practices and community nursing services to obtain a maximum diversity sample of team cultures and practices. (28,29,53) Rich descriptions of practice aid understanding and transferability of our results. (53) Our methods enabled detailed qualitative and quantitative analysis of recorded events and their context, which would not have been possible through analysis of the available large primary care datasets. (26,49,54) AMs prescribing data, context and decision-making are not routinely recorded in a way which is systematically retrievable by using electronic algorithms. Consequently, details of care in the body of free text records are often overlooked in large database studies and valuable insights into practice are missed. (26,54)

We collected complete data from patient electronic clinical records: five patient community nursing paper prescription charts were missing. Prescribing events and contexts were confirmable from electronic records, and we present a full data set for all the variables analysed. As clinical records are not designed to collect research data, some patient characteristics were not routinely recorded, such as cohabitation status or perceived risks of opioids being misused or diverted, factors that may influence AM prescribing (20). Data concerning the administration of AMs for these patients will be presented in a forthcoming paper.

## Conclusions

This mixed-methods clinical records study provides valuable insights into an important area of community end-of-life care practice. Standardised AMs prescribing patterns suggest undue reliance on electronic end-of-life care templates and a lack of individualised prescribing as advocated in national policy. Marked variability in the timing of prescriptions, at times many months before death, underscores the challenge of prognostication and highlights the risks involved in putting medication in place too far in advance of possible need. The presence of AMs for long periods of time, or when situations are uncertain, may therefore compromise patient safety unless robust systems are in place to review their continued appropriateness and safe use.

## Supporting information

Supplementary Document 2 - Multivariable Logistic Regression

## Data Availability

Data sharing: The anonymised prescribing quantitative data used in this study may be requested by researchers on completion of a data use agreement.

## Acknowledgements

The authors would like to thank the Positive Ageing PPI Group, Cambridge Palliative and End of Life Care PPI Group for their helping planning the study and interpreting the findings. James Brimicombe, Senior Data Manager for the Primary Care Unit, Department of Public Health and Primary, University of Cambridge, for his help in planning and running this study. Angela Harper for her administrative support. Efthalia (Lina) Massou, Statistician and Research Associate, for her statistical support. The National Institute for Health Research Clinical Research Network (NIHR CRN) for their study support. Hertfordshire Community NHS Trust and the other research sites for providing access to patient records.

## Declaration of conflict of interests

The authors declare no potential conflicts of interest

## Funding

BB is funded by the National Institute for Health Research (NIHR) School for Primary Care Research. SB is funded by the NIHR Applied Research Collaboration East of England (ARC EoE) programme. This work was also supported by the RCN Foundation Professional Bursary Scheme [grant number 20181113]

The views expressed are those of the author(s) and not necessarily those of the NIHR or the Department of Health and Social Care.

## Contributors

BB, KP and SB designed the study. BB carried out the analysis with input from KP and SB. All authors contributed to the interpretation of the results and the writing of the manuscript. BB is the guarantor. The corresponding author attests that all listed authors meet authorship criteria and that no others meeting the criteria have been omitted. The lead author (BB) affirms that the manuscript is an honest accurate, and transparent account of the study being reported.

## Data sharing

The anonymised prescribing quantitative data used in this study may be requested by researchers on completion of a data use agreement at (Repository URL to be confirmed on acceptance).

## Notes

### Competing Interest Statement

The authors have declared no competing interest.

### Author Declarations

The South Cambridgeshire Research Ethics Committee granted ethics approval [Reference: 19/EE/0012]. The Health Research Authority Confidentiality Advisory Group [19/CAG/0014] approved the processing of confidential patient information without consent in the public interest: data were anonymised at the earliest opportunity.

