## Supplementary Document 2 - Multivariable Logistic Regression for "Unwelcome memento mori or best clinical practice? Community end-of-life anticipatory medication prescribing practice: a mixed methods observational study"

| **Supplementary Table 1. Prescribing of anticipatory medication: Multivariable Logistic Regression** | | | |  |  |  |  |  |
| --- | --- | --- | --- | --- | --- | --- | --- | --- |
|  |  |  |  |  |  |  | 95% CI for Exp (B) | |
| Step 3a. | B | SE | Wald | df | Sig. | Exp (B) | Lower | Upper |
| **Gender** (female vs. male) | -0.091 | 0.382 | 0.057 | 1 | 0.811 | 0.913 | 0.431 | 1.931 |
| **Age Range** (vs. 85+), years |  |  | 2.426 | 3 | 0.489 |  |  |  |
| 18-64 | -1.005 | 0.657 | 2.342 | 1 | 0.126 | 0.366 | 0.101 | 1.326 |
| 65-74 | -0.505 | 0.625 | 0.652 | 1 | 0.419 | 0.603 | 0.177 | 2.056 |
| 75-84 | -0.213 | 0.511 | 0.174 | 1 | 0.677 | 0.808 | 0.297 | 2.199 |
| **GP Practice ID No.**  Reference = No. Eleven |  |  | 12.366 | 10 | 0.261 |  |  |  |
| One | 0.19 | 0.903 | 0.044 | 1 | 0.833 | 1.209 | 0.206 | 7.102 |
| Two | 0.554 | 0.973 | 0.324 | 1 | 0.569 | 1.74 | 0.259 | 11.714 |
| Three | 0.193 | 0.917 | 0.044 | 1 | 0.833 | 1.213 | 0.201 | 7.32 |
| Four | 2.632 | 1.099 | 5.738 | 1 | 0.017 | 13.9 | 1.614 | 119.75 |
| Five | 0.812 | 0.919 | 0.78 | 1 | 0.377 | 2.252 | 0.372 | 13.634 |
| Six | -0.288 | 0.891 | 0.104 | 1 | 0.747 | 0.75 | 0.131 | 4.302 |
| Seven | 0.659 | 0.898 | 0.539 | 1 | 0.463 | 1.934 | 0.332 | 11.249 |
| Eight | 1.093 | 0.965 | 1.283 | 1 | 0.257 | 2.982 | 0.45 | 19.764 |
| Nine | 1.03 | 0.99 | 1.083 | 1 | 0.298 | 2.802 | 0.402 | 19.515 |
| Ten | -0.345 | 0.996 | 0.12 | 1 | 0.729 | 0.708 | 0.101 | 4.986 |
| **Number of chronic disease**  **registers on** (vs. 8-13) |  |  | 1.745 | 4 | 0.783 |  |  |  |
| 0-1 | -0.394 | 0.886 | 0.197 | 1 | 0.657 | 0.674 | 0.119 | 3.833 |
| 2-3 | -0.225 | 0.715 | 0.099 | 1 | 0.753 | 0.799 | 0.197 | 3.244 |
| 4-5 | -0.339 | 0.726 | 0.218 | 1 | 0.64 | 0.712 | 0.172 | 2.954 |
| 6-7 | 0.474 | 0.843 | 0.317 | 1 | 0.574 | 1.607 | 0.308 | 8.378 |
| **Usual Place of Residence**  (care home vs. home) | -0.023 | 0.699 | 0.001 | 1 | 0.974 | 0.977 | 0.248 | 3.842 |
| **Cause of Death** (cancer vs. non-cancer) | 0.27 | 0.517 | 0.273 | 1 | 0.601 | 1.31 | 0.475 | 3.61 |
| **Preferred Place of Death** (recorded vs. not recorded) | 3.533 | 0.416 | 72.266 | 1 | 0.000 | 34.238 | 15.16 | 77.322 |
| **Received Specialist Palliative Care** (yes vs. no) | 1.96 | 0.492 | 15.902 | 1 | 0.000 | 7.101 | 2.71 | 18.611 |
| Constant | -3.028 | 1.107 | 7.49 | 1 | 0.006 | 0.048 |  |  |
| a. Variables entered: Gender, Age Range, GP Practice, Number Chronic Disease Register Groups, Usual Residence, | | | | | | |  |  |
| **Cause of Death, Preferred Place of Death, Seen Specialist Palliative Care.**  **Steps 1-3 are detailed in the Supplemental Document 3.** | | | | | | |  |  |
